# Using Artificial Intelligence for COVID-19 Chest X-ray Diagnosis

**DOI:** 10.1101/2020.05.21.20106518

**Authors:** Andrew A. Borkowski, Narayan A. Viswanadhan, L. Brannon Thomas, Rodney D. Guzman, Lauren A. Deland, Stephen M. Mastorides

**Affiliations:** Pathology and Laboratory Medicine Service, James A. Haley Veterans’ Hospital, Tampa, Florida, USA.; Radiology Service, James A. Haley Veterans’ Hospital, Tampa, Florida, USA.; Department of Pathology and Cell Biology, University of South Florida, Morsani College of Medicine, Tampa, Florida, USA.; InterKnowlogy, LLC, Carlsbad, California, USA.

**Keywords:** COVID-19, coronavirus, AI, diagnosis, radiology, pathology, deep learning

## Abstract

Coronavirus disease-19 (COVID-19), caused by a novel member of the coronavirus family, is a respiratory disease that rapidly reached pandemic proportions with high morbidity and mortality. It has had a dramatic impact on society and world economies in only a few months. COVID-19 presents numerous challenges to all aspects of healthcare, including reliable methods for diagnosis, treatment, and prevention. Initial efforts to contain the spread of the virus were hampered by the time required to develop reliable diagnostic methods. Artificial intelligence (AI) is a rapidly growing field of computer science with many applications to healthcare. Machine learning is a subset of AI that employs deep learning with neural network algorithms. It can recognize patterns and achieve complex computational tasks often far quicker and with increased precision than humans. In this manuscript, we explore the potential for a simple and widely available test as a chest x-ray (CXR) to be utilized with AI to diagnose COVID-19 reliably. Microsoft CustomVision is an automated image classification and object detection system that is a part of Microsoft Azure Cognitive Services. We utilized publicly available CXR images for patients with COVID-19 pneumonia, pneumonia from other etiologies, and normal CXRs as a dataset to train Microsoft CustomVision. Our trained model overall demonstrated 92.9% sensitivity (recall) and positive predictive value (precision), with results for each label showing sensitivity and positive predictive value at 94.8% and 98.9% for COVID-19 pneumonia, 89% and 91.8% for non-COVID-19 pneumonia, 95% and 88.8% for normal lung. We then validated the program using CXRs of patients from our institution with confirmed COVID-19 diagnoses along with non-COVID-19 pneumonia and normal CXRs. Our model performed with 100% sensitivity, 95% specificity, 97% accuracy, 91% positive predictive value, and 100% negative predictive value. Finally, we developed and described a publicly available website to demonstrate how this technology can be made readily available in the future.

## 1. INTRODUCTION

The novel coronavirus Severe Acute Respiratory Syndrome coronavirus 2 (SARS-COV-2), which causes the respiratory disease Coronavirus disease-19 (COVID-19), was first identified as a cluster of cases of pneumonia in Wuhan, Hubei Province of China on December 31, 2019.^1^ Within a month, the disease had spread significantly, leading the World Health Organization (WHO) to designate COVID-19, a Public Health Emergency of International Concern (PHEIC). On March 11, 2020, the WHO declared COVID-19 a global pandemic.^2^ As of May 7, 2020, the virus has infected more than 3.6 million people, with over 250,000 deaths worldwide.^3^ The dramatic impact the spread of COVID-19 has had on social, economic, and healthcare issues throughout the world has been reviewed.^4^

Prior to the 21st century, members of the coronavirus family had minimal impact on human health.^5^ However, in the past 20 years, outbreaks have highlighted an emerging importance of coronaviruses in morbidity and mortality on a global scale. Although less prevalent than COVID-19, severe acute respiratory syndrome (SARS) in 2002-2003 and Middle Eastern respiratory syndrome (MERS) in 2012 likely had higher mortality rates than the current pandemic.^5^ Based on this recent history, it is reasonable to assume that we will continue to see novel diseases with similar significant health and societal implications. The challenges presented to health care providers by such novel viral pathogens are numerous, including methods for rapid diagnosis, prevention, and treatment. In the current study, we focus on diagnosis issues, which were evident with COVID19 with the time required to develop rapid and effective diagnostic modalities.

We have previously reported the utility of using artificial intelligence (AI) in the histopathologic diagnosis of cancer.^6–8^ AI was first described in 1956 and involves the field of computer science in which machines are trained to learn from experience.^9^ Machine learning (ML) is a subset of AI and is achieved by using mathematical models to compute sample data sets.^10^ Current ML utilization employs deep learning with neural networks algorithms, which can recognize patterns and achieve complex computational tasks often far quicker and with increased precision than can humans.^11–13^ In addition to applications in pathology, ML algorithms have both prognostic and diagnostic applications in multiple medical specialties such as radiology, dermatology, ophthalmology, and cardiology.^6^ It is predicted that AI will impact almost every aspect of health care in the future.^14^

In this manuscript, we examine the potential for AI to diagnose patients with COVID-19 pneumonia using chest radiographs (CXR) alone. This is done using Microsoft CustomVision, a readily available, automated ML platform.^15^ Employing AI to both screen and diagnose emerging health emergencies such as COVID-19 has the potential to dramatically change how we approach medical care in the future. In addition, we describe the creation of a publicly available website that could augment COVID-19 pneumonia CXR diagnosis.

## 2. MATERIALS AND METHODS

### 2.1 Training Dataset

One hundred three CXR images of COVID-19 were downloaded from GitHub covid-chest-xray dataset.^16^ Five hundred images of non-COVID-19 pneumonia and 500 images of the normal lung were downloaded from the Kaggle RSNA Pneumonia Detection Challenge dataset.^17^ To balance the dataset, we expanded the COVID-19 dataset to 500 images by slight rotation (probability=1, max rotation=5) and zooming (probability=0.5, percentage_area=0.9) of the original images using the Augmentor python package.^18^

### 2.2 Validation Dataset

Thirty random CXR images from the Veteran's Administration (VA) PAC system were obtained for the validation dataset. This dataset included ten CXR images from hospitalized COVID-19 patients, ten CXR pneumonia images from non-COVID-19 patients, and ten normal CXRs. COVID-19 diagnoses were confirmed with a positive test result from the Xpert® Xpress SARS-CoV-2 PCR platform.^19^

### 2.3 Microsoft CustomVision

Microsoft CustomVision is an automated image classification and object detection system that is a part of Microsoft Azure Cognitive Services.^15^ It has a pay as you go model with fees depending on your computing needs and usage. It offers a free trial to users for two initial projects. The service is web-based with an easy to follow graphical user interface. No coding skills are necessary.

### 2.4 Model Training

In Microsoft CustomVision, we created a new project with the following setup: project type – classification, classification type – multiclass (single tag per image), domains – compact general for small size and easy export to TensorFlow.js model format. With the project created, we proceeded to upload our image dataset. Each class was uploaded separately and tagged with the appropriate label (covid_pneumonia, non-covid pneumonia, and normal lung). The system rejected 16 COVID-19 images as duplicates. The final CustomVision training dataset consisted of 484 images of COVID-19 pneumonia, 500 images of non-COVID-19 pneumonia, and 500 images of normal lung. Once uploaded, Microsoft CustomVision self-trains using the dataset upon initiating the program. (Figure 1)

**Figure 1.**
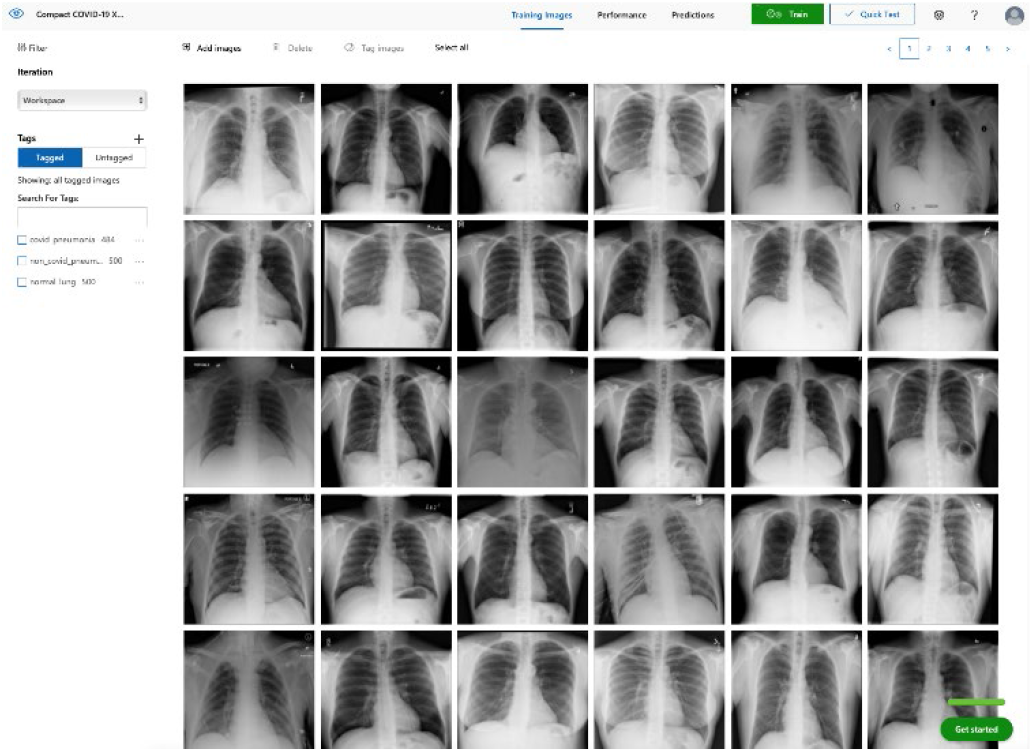
Screenshot of image datasets uploaded to MS CustomVision.

### 2.5 Website Creation

Microsoft Azure Custom Vision was used to train the model. Custom Vision can be used continuously to execute the model, or the model can be compacted and decoupled from Azure. In this case, the model was compacted and decoupled for use in a web app. An Angular web app was created with TensorFlow.js. TensorFlow.js is a JavaScript library that enables dynamic download and execution of ML models. Within a user's web browser, the model is executed when an image of a CXR is submitted. Confidence values for each classification are returned. In this design, after the initial web page and model is downloaded, the web page no longer needs to access any server components and performs all operations in the browser. Although the solution works well on mobile phone browsers and in low bandwidth situations, the quality of predictions may depend on the browser and device one uses. At no time does an image get submitted to the cloud.

## 3. RESULTS

Overall, our trained model showed 92.9% precision and recall. Precision and recall results for each individual label were 98.9% and 94.8% for COVID-19 pneumonia, 91.8% and 89% for non-COVID-19 pneumonia, and 88.8% and 95% for normal lung. (Figure 2) Next, we proceeded to validate the training model on the VA data by making individual predictions on 30 images from the VA dataset. Our model performed very well with 100% sensitivity (recall), 95% specificity, 97% accuracy, 91% positive predictive value (precision), and 100% negative predictive value.

**Figure 2.**
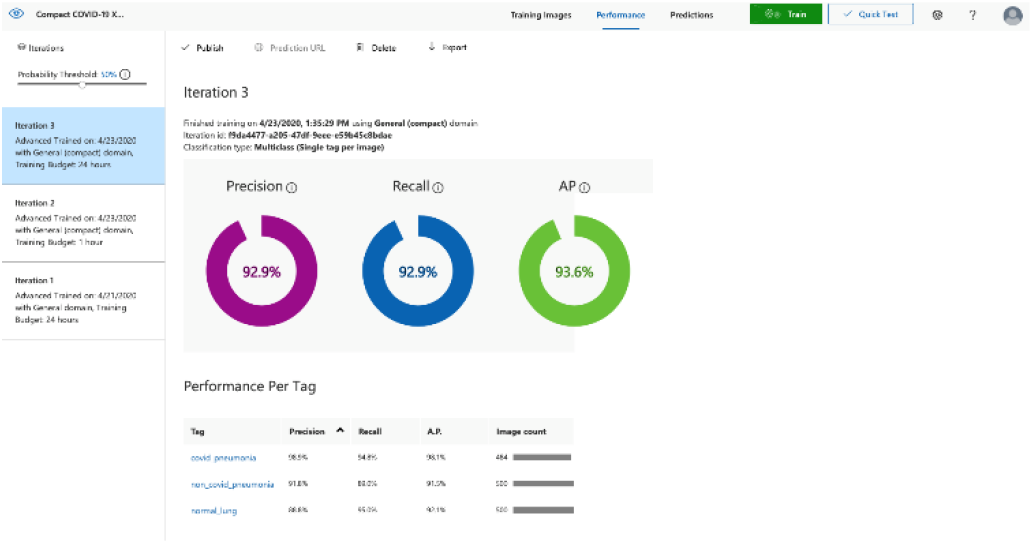
Screenshot of the trained ML model performance.

## 4. DISCUSSION

We have successfully demonstrated the potential of using AI algorithms in assessing CXRs for COVID-19. We first trained the Microsoft CustomVision automated image classification and object detection system to differentiate cases of COVID- 19 from pneumonia from other etiologies as well as normal lung CXRs. We then tested our model against known patients from our medical center. The program achieved 100% sensitivity (recall), 95% specificity, 97% accuracy, 91% positive predictive value (precision), and 100% negative predictive value in differentiating the three scenarios. Using the trained ML model, we proceeded to create a website that could augment COVID-19 CXR diagnosis.^20^ The website works on mobile as well as desktop platforms. One can take a CXR photo with a mobile phone or upload it from the file. The ML algorithm would provide the probability of COVID-19 pneumonia, non-COVID-19 pneumonia, or normal lung X-ray diagnosis. (Figure 3)

**Figure 3.**
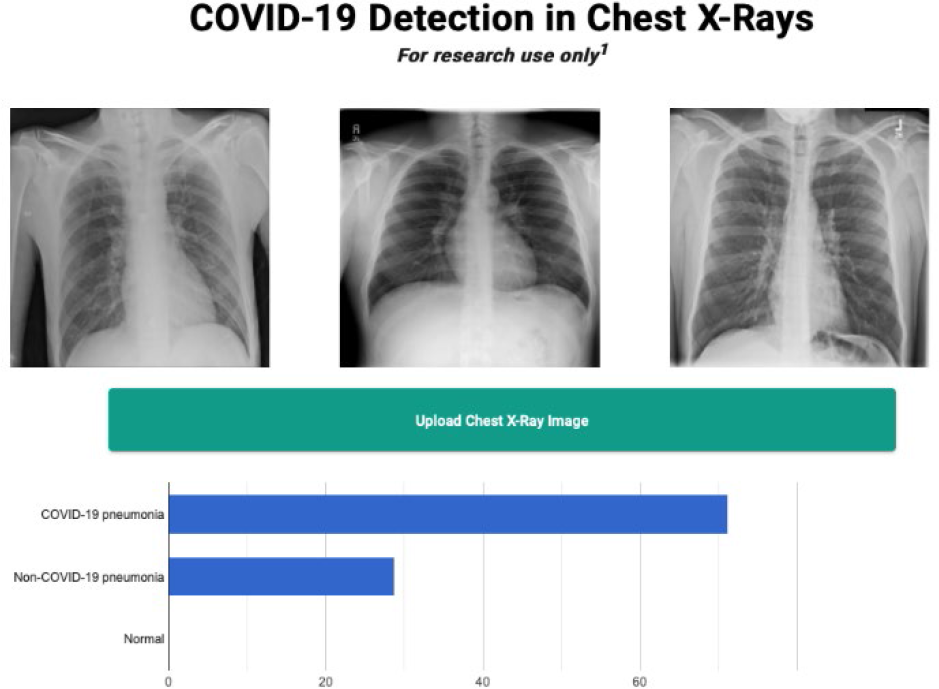
Screenshot of the website.

Emerging diseases such as COVID-19 present numerous challenges to healthcare providers, governments, and businesses, as well as to individual members of society. As evidenced with COVID-19, the time from first recognition of an emerging pathogen to the development of methods for reliable diagnosis and treatment can be months, even with a concerted international effort. The gold standard for diagnosis of COVID-19 is by reverse transcriptase polymerase chain reaction (RT-PCR) technologies, but early RT-PCR testing produced less than optimal results.^21–23^ Even after the development of reliable tests for detection, making test kits readily available to health care providers on an adequate scale presents an additional challenge as evident with COVID-19.

The lack of availability of diagnostic RT-PCR with COVID- 19 initially placed increased reliability on presumptive diagnoses via imaging in some situations.^24^ Most of the literature evaluating radiographs of COVID-19 patients focuses on chest computed tomography (CT) findings, with initial results suggesting CT was more accurate than early RT-PCR methodologies.^22,23,25^ The Radiological Society of North America Expert consensus statement on chest CT for COVID- 19 states that CT findings can even precede positivity on RT-PCR in some cases.^23^ However, currently they do not recommend the use of CT scanning as a screening tool. Furthermore, the actual sensitivity and specificity of CT interpretation by radiologists for COVID-19 are unknown.^23^

Characteristic CT findings include ground-glass opacities (GGOs) and consolidation most commonly in the lung periphery, though a diffuse distribution was found in a minority of patients.^22,24,26–28^ Lomoro and colleagues recently summarized the CT findings from several reports which described abnormalities as most often bilateral and peripheral, subpleural and affecting the lower lobes.^27^ Not surprisingly, CT appears more sensitive at detecting changes with COVID- 19 than CXR, with reports that a minority of patients had CT changes before changes visible on CXR.^24,27^

We focused our study on the potential of AI in the examination of CXRs in patients with COVID-19, as there are several limitations to the routine use of CT scans with conditions such as COVID-19. Aside from the more considerable time required to obtain CTs, there are issues with contamination of CT suites, sometimes requiring a dedicated COVID-19 CT scanner.^24,29^ The time constraints of decontamination or limited utilization of CT suites can delay or disrupt services for both COVID-19 and non-COVID-19 patients. Because of these factors, CXR may be a better resource to minimize the risk of infection to other patients. Besides, accurate assessment of abnormalities on CXR for COVID-19 may identify patients in whom the CXR was performed for other purposes.^24^ CXR is more readily available than CT, especially in more remote or underdeveloped areas.^29^ Finally, as with CT, CXR abnormalities are reported to have appeared before RT-PCR tests became positive in a minority of patients.^24^

CXR findings described in COVID-19 patients are similar to those of CT and include GGOs, consolidation, and hazy increased opacities.^24,26,27,29,30^ Like CT, the majority of patients demonstrated greater involvement in the lower zones and peripherally^24,26,27,29,30^ Most patients showed bilateral involvement. However, while these findings are common in COVID-19 patients, they are not specific and can be seen in other conditions such as other viral pneumonia, bacterial pneumonia, injury from drug toxicity, inhalation injury, connective tissue disease, and idiopathic conditions.

Applications of AI in interpreting radiographs of various types are numerous, and extensive literature has been written on the topic.^31^ Using deep learning algorithms, AI has multiple possible roles to augment traditional radiograph interpretation. These include the potential for screening, triaging, and increasing the speed to render diagnoses. It also can provide a rapid “second opinion” to the radiologist to support the final interpretation. In areas with critical shortages of radiologists, it potentially can be used to render the definitive diagnosis. With COVID-19, imaging studies have been shown to correlate with disease severity and mortality, and AI could assist in monitoring the course of the disease as it progresses and potentially identifies patients at greatest risk.^28^ There is excellent potential should a rapid diagnostic test as simple as a CXR be able to reliably impact containment and prevention of the spread of contagions such as COVID-19 early in its course.

Few studies have assessed using AI in the radiologic diagnosis of COVID-19, most of which utilize CT scanning. Bai and colleagues demonstrated increased accuracy, sensitivity, and specificity in distinguishing chest CTs of COVID-19 patients from other types of pneumonia.^22,32^ A separate study demonstrated the utility of using AI to differentiate COVID- 19 from community-acquired pneumonia with CT.^33^ However, the effective utility of AI for CXR interpretation has been demonstrated as well.^14,34^ Implementation of convolutional neural network layers has allowed for reliable differentiation of viral and bacterial pneumonia with CXR imaging.^35^ Evidence suggests that there is great potential in the application of AI in the interpretation of radiographs of all types.

Finally, as mentioned, we have developed a publicly available website based on our studies.^20^ It should be stressed that this website is for research use only. To utilize the website, images must have protected health information (PHI) removed before uploading. The information on the website, including texts, graphics, images, or other material, is for research purposes and may not be appropriate for all circumstances. The website does not provide medical, professional, or licensed advice and is not a substitute for consultation with a health care professional. Medical advice should be sought from a qualified health care professional for any questions, and the website should not be used for medical diagnosis or treatment.

## 5. CONCLUSIONS

We have utilized a readily available, commercial platform to demonstrate the potential of AI to assist in the successful diagnosis of COVID-19 pneumonia on CXR images. While this technology has numerous applications in radiology, we have focused on the potential impact on future world health crises such as COVID-19. The findings have implications for screening and triage, initial diagnosis, monitoring disease progression, and identifying patients at increased risk of morbidity and mortality. Based on the data, a website was created to demonstrate how such technologies could be shared and distributed to others to combat entities such as COVID-19 moving forward. Our study offers a small window into the potential for how AI will likely dramatically change the practice of medicine in the future.

## Data Availability

Publicly available images were utilized in this study from the following two websites.

https://arxiv.org/abs/2003.11597

https://www.kaggle.com/datasets

## ACKNOWLEDGMENTS

None

## FUNDING AND ETHICAL DISCLOSURE

This material is the result of work supported with resources and the use of facilities at the James A. Haley Veterans’ Hospital and computational resources of the InterKnowlogy, LLC. This activity has been approved by the James A. Haley Veterans’ Hospital Office of Research and Development.

